# Heterogeneity of hypoxemia severity according to oxygenation index in COVID-19 pneumonia

**DOI:** 10.1101/2021.06.23.21259421

**Authors:** Isaac Núñez, Adrian Soto-Mota

**Author notes:** **Corresponding author:** Isaac Núñez MD, Department of Internal Medicine, Instituto Nacional de Ciencias Médicas y Nutrición Salvador Zubirán, Mexico City, Mexico. Vasco de Quiroga # 15, Belisario Domínguez Sección XVI, Tlalpan, Mexico City, 14080, Mexico. The authors will not ask for reprints.

## Abstract

**Objective:** To compare hypoxemia severity of patients with COVID-19 pneumonia that arrive at an emergency department as classified by three oxygenation indexes.

**Design:** Retrospective analysis of pulse oximeter saturation and arterial blood gas analysis obtained at arrival.

**Setting:** Tertiary referral hospital in Mexico City converted early in the pandemic to a COVID-19 center.

**Patients and measurements:** A total of 2,960 patients with suspected COVID-19 pneumonia were admitted to the emergency department from April 2020 until March 2021. Pulse oximeter saturation and arterial blood gas analysis was obtained in all of them. Pulse oximeter saturation (SpO2) to inspired oxygen fraction ratio (FiO2), oxygen saturation in arterial blood (SatO2) to FiO2 ratio, and oxygen pressure in arterial blood to FiO2 ratio were calculated for every patient.

**Interventions:** None.

**Main Results:** A strong correlation was seen between PaO2/FiO2 & SpO2/FiO2 (rho = 0.6, p < 0.001), and SatO2/FiO2 & SpO2/FiO2 (rho = 0.65, p < 0.001), while a very strong correlation was seen between PaO2/FiO2 & SatO2/FiO2 (rho = 0.88, p < 0.001). When classifying severity by quantiles, considerable cross-over was observed when comparing oxygenation indexes, as only 785 (26.5%) patients were in the same quintile across the three indexes.

**Conclusions:** Hypoxemia severity is heterogeneous according to the oxygenation index utilized. This limits their usefulness as sole markers of severity, as inter-observer variability, especially on FiO2 estimation, and different practices limit consistent follow up and treatment decisions.

## INTRODUCTION

Pneumonia is the hallmark of severe COVID-19 (1). Strain in healthcare systems across the world has forced countless hospitals to conduct grueling triages to decide who gets to be admitted when healthcare saturation was rampant (2). As these decisions are inherently complex, numerous risk scores and predictor factors have been described to aid the attending medical team (3–5). These often include clinical and laboratory values.

One commonly utilized criteria to determine patient severity is the severity of hypoxia (6). This can be assessed with arterial oxygen pressure (PaO2), PaO2 to oxygen inspired fraction (FiO2) ratio, arterial oxygen saturation (SatO2), pulse oximeter oxygen saturation (SpO2), SatO2 to FiO2 ratio, SpO2 to FiO2 ratio, and the prescribed oxygen device (for example, nasal cannula, non-rebreather mask, Venturi mask, etc) (7).

Recently, the use of these criteria for hypoxia severity in non-intubated patients has been criticized given the expected high inter-patient variability in FiO2, shunt fraction, and physician’s choice of oxygenation device and oxygen flow (7). Therefore, relying on these criteria is suboptimal given the low comparability between different patients.

In this study, we aimed to compare the severity of hypoxemia in COVID-19 patients with severe pneumonia arriving at an emergency department.

## METHODS

### Study site description

We performed a retrospective cohort study collecting information on every patient who arrived at the emergency department (ED) of a large COVID-19 tertiary center in Mexico City between April 1^st,^ 2020, and April 30^th^, 2021. At arrival, every patient had to go through a triage station where vital signs (including SpO2) were documented before entering the emergency department. Once in the emergency department, all patients who had low SpO2 (defined as < 92%) received supplemental oxygen. Only nasal cannula and non-rebreathing masks were available at our center. Arterial blood gas analysis was performed in all patients with suspected COVID-19 and low SpO2. Generally, FiO2 was estimated heuristically by the treating physician by adding to the baseline FiO_2_ (21%) 3% for every extra liter of oxygen per minute (L/min). For example, a patient receiving 2 L/min of supplemental oxygen would have a calculated FiO2 of 27% (21 + 3*2). SpO2 was obtained at ambient air, while arterial blood gas was obtained almost universally when patients received supplemental oxygen. Given the closeness between SpO2 and blood gas analysis, we believe it is reasonable to assume that the clinical status of the patient is comparable between these two circumstances. In case more than one emergency episode was documented for a given individual, only the first one in which an arterial blood gas was obtained was included.

### Statistical analysis

We obtained data about PaO2, SatO2, SpO2, and FiO2, with which we calculated PaO2/FiO2 and SatO2/FiO2. All SpO2 were taken at triage, and as such, were ambient-air (FiO2 21%). SpO2/FiO2 was included to better compare oxygenation indexes. The oxygen device used at the time of the arterial blood gas analysis could not be confidently determined, so it was not included. Since the low reliability of FiO2 is the most criticized aspect of using the oxygenation device as a marker of hypoxia severity, it does not affect our analysis.

We categorized a patient’s hypoxemia severity by quintiles of SpO2/FiO2, PaO2/FiO2, and SatO2/FiO2 (from now on referred to as “oxygenation indexes”). Lower values indicate a higher hypoxemia severity. We determined the strength of concordance between oxygenation index quintile category with chord diagrams across all three oxygenation index pairs. We calculated Spearman correlation coefficients for the three possible pairs of oxygenation indexes. We built scatterplots and used locally weighted scatterplot smoothing regression to graphically represent the data.

All analyses were conducted with R software version 4.0.0. The study was approved by the ethics committee of the Instituto Nacional de Ciencias Médicas y Nutrición Salvador Zubirán.

## RESULTS

A total of 23,049 triage visits occurred during the study period, corresponding to 19,644 individual patients. Of these, 8,123 had a visit due to suspected COVID-19. Among patients that were finally admitted to the ED, necessary data could be obtained for 3,013 patients. Only 53 patients were excluded due to implausibly high PO2 levels (>100 with FiO2 of 21%). Thus, 2,960 patients were finally included in the analysis.

Median FiO2 was 0.34 (inter-quartile range [IQR] 0.25-0.60), PaO2 68 mmHg (57-84), SatO2 94% (91-97), SpO2 83% (73-88), PaO2/FiO2 211 (124-281), SatO2/FiO2 274 (160-364), and SpO2/FiO2 395 (348-419). Correlation among oxygenation indexes is shown in **Figure 1**. A strong correlation was seen between PaO2/FiO2 & SpO2/FiO2 (rho = 0.6, p < 0.001), and SatO2/FiO2 & SpO2/FiO2 (rho = 0.65, p < 0.001), while a very strong correlation was seen between PaO2/FiO2 & SatO2/FiO2 (rho = 0.88, p < 0.001). None of the variable pairs showed a linear relationship. All oxygenation indexes showed a considerable cross-over among quintiles (**Figure 2**), with 785 (26.5%) patients being in the same severity quintile across all indexes.

**Figure 1.**
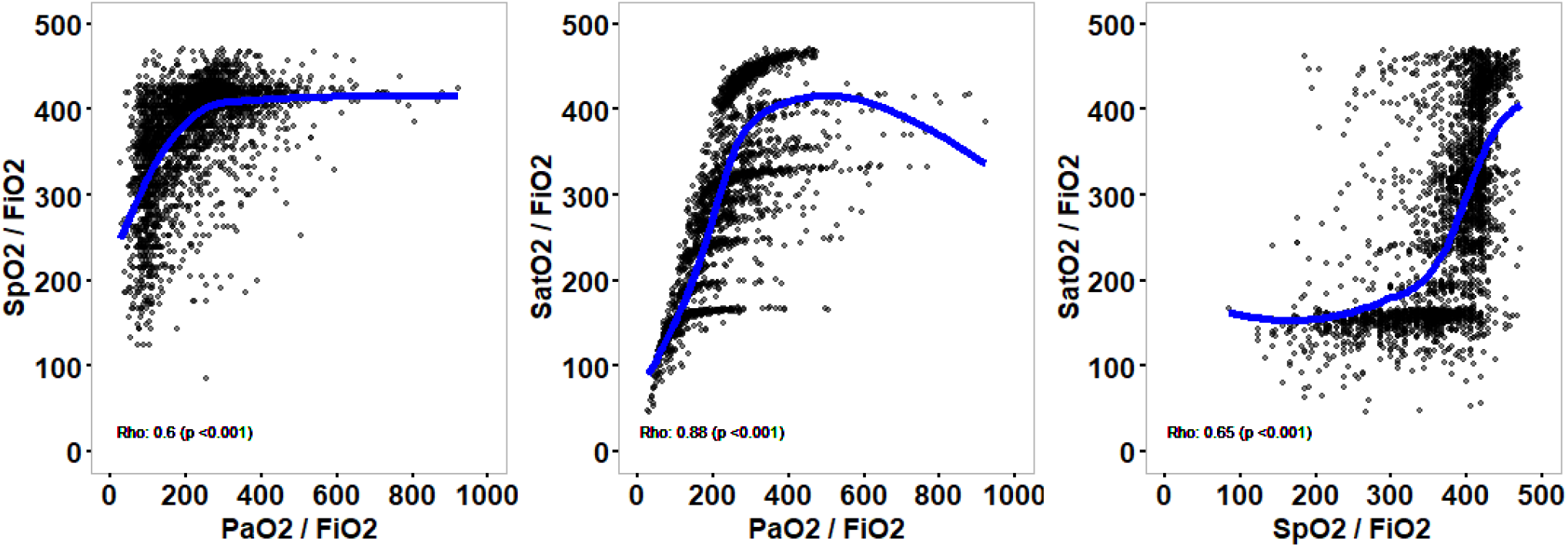
Correlation between oxygenation indexes. SpO2: Oxygen saturation with pulse oximeter; PaO2: oxygen pressure in arterial blood; SatO2: oxygen saturation in arterial blood; FiO2: fraction of inspired oxygen.

**Figure 2.**
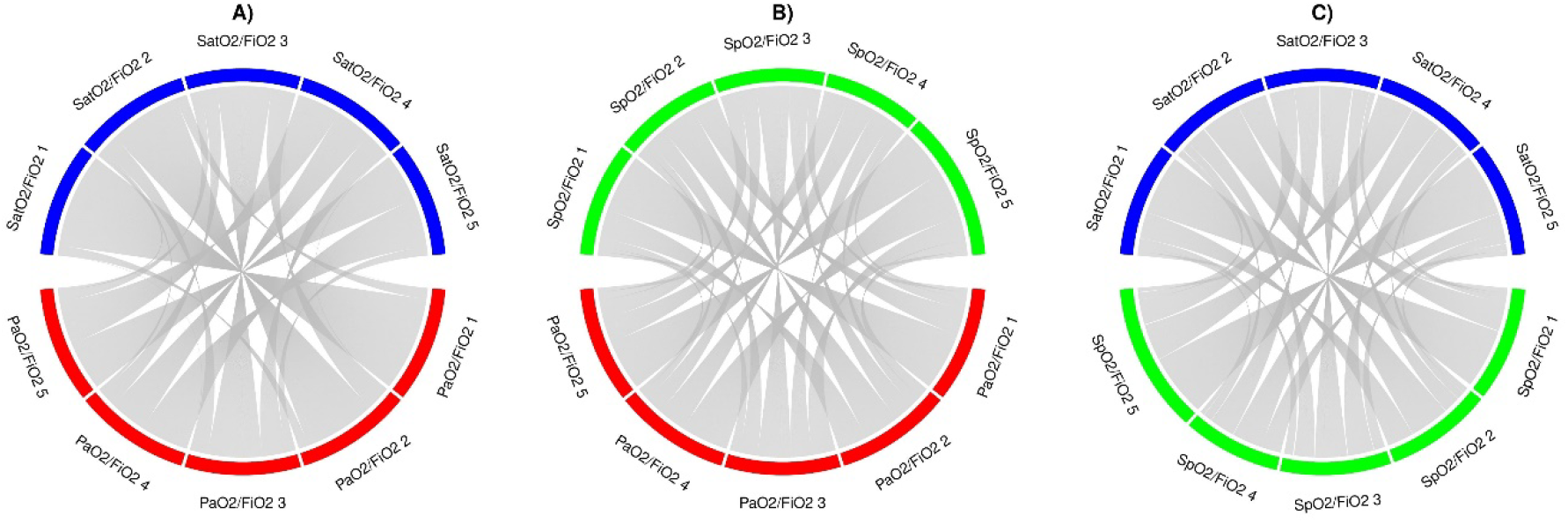
Strength of relation between oxygenation indexes by quintiles. SpO2: Oxygen saturation with pulse oximeter; PaO2: oxygen pressure in arterial blood; SatO2: oxygen saturation in arterial blood; FiO2: fraction of inspired oxygen. A) Shows relation between PaO2/FiO2 and SatO2/FiO2, B) shows relation between PaO2/FiO2 and SpO2/FiO2, and C) shows relation between SpO2/FiO2 and SatO2/FiO2. Numbers represent the respective quintile (“PaO2/FiO2 1” represents the first quintile of said variable).

## DISCUSSION

Our study shows there is considerable heterogeneity when classifying hypoxemia severity with different oxygenation indexes. While good correlation was observed among the three evaluated pairs, the lower correlation between SpO2/FiO2 and the others could be due to inaccuracy of pulse oximeters when used in patients with low oxygen saturation, physician imprecision when estimating FiO2, and/or mixed blood in the gas analysis (8). It is likely multiple factors are involved in most cases, which is consistent with the previously mentioned concerns (7). For example, if solely based on SpO2 at triage, a patient could be wrongly classified as having a more severe disease, giving preference to a patient in better condition. Also, follow up could be hard if it is done only with oxygenation parameters. The morning medical team could classify the patient with a given severity with blood gas analysis, while the evening group could use a pulse oximeter.

Overall, our study adds to the argument that oxygenation is not a good measure by itself in COVID-19 pneumonia, as considerable variation when classifying disease severity is likely.

## Data Availability

Code used for the analysis is available at https://github.com/isaac-nunez/Oxygenation_indexes.

## Data availability

Code used for the analysis is available at https://github.com/isaac-nunez/Oxygenation_indexes.

